# *In vivo* delayed clearance of *Plasmodium falciparum* malaria independent of *kelch13* polymorphisms and with escalating malaria in Bangladesh

**DOI:** 10.1101/2021.06.24.21259123

**Authors:** Maisha Khair Nima, Saiful Arefeen Sazed, Angana Mukherjee, Muhammad Riadul Haque Hossainey, Ching Swe Phru, Fatema Tuj Johora, Innocent Safeukui, Anjan Saha, Afsana Alamgir Khan, Aung Swi Prue Marma, Russell E. Ware, Mohandas Narla, Barbara Calhoun, Rashidul Haque, Wasif Ali Khan, Mohammad Shafiul Alam, Kasturi Haldar

**Affiliations:** Boler-Parseghian Center for Rare and Neglected Diseases, Dept. of Biological Sciences, University of Notre Dame, Notre Dame IN 46556, USA; Infectious Diseases Division, International Centre for Diarrhoeal Disease Research, Bangladesh (icddr,b), Mohakhali, Dhaka 1212, Bangladesh; Department of Biological Sciences, George Washington University, Washington, DC 20052, USA; National Malaria Elimination & Aedes Transmitted Diseases Control Program, DGHS, Mohakhali, Dhaka 1212, Bangladesh; Civil Surgeon’s Office, Bandarban, Bangladesh; Division of Hematology, Department of Pediatrics, and the Global Health Center, Cincinnati Children’s Hospital Medical Center, Cincinnati OH 45229, USA; New York Blood Center, New York, NY 10065, USA

## Abstract

The emergence of resistance to artemisinin drugs threatens global malaria control. Resistance is widely seen in South East Asia (SEA) and Myanmar, but not comprehensively assessed in Bangladesh. This is due to lack of measuring parasite clearance times in response to drug treatment, a gold standard used to track artemisinin resistance (AR), in the Chittagong Hill Tracts (CHT), where >90% of malaria occurs in Bangladesh. Here we report delay in clinical parasite clearance half-lives > 5 h characteristic of AR, in Bandarban, a south-eastern rural, CHT district with escalating malaria and bordering Myanmar. Thirty-one and 68 malaria patients respectively presented in the clinic in 2018 and 2019, and this increase well correlated to the district-level malaria surge and rise in rainfall, humidity and temperature. A total of 27 patients with uncomplicated *Plasmodium falciparum* malaria mono-infection, after administration of an artemisinin combination therapy (ACT) showed median (range) parasite clearance half-life and time of 5.6 (1.5 – 9.6) and 24 (12–48) hours (h) respectively. The frequency distribution of parasite clearance half-life and time was bimodal, with a slower parasite clearance of 8 h in 20% of the participants. There was however, no detectable parasitemia 72 h after initiating ACT. Half-life clearance of > 5h, respectively seen in 35% and 40% of participants in 2018 and 2019, lacked in correlation to initial parasitemia, blood count parameters or resistance mutations of *PfKelch13* (K13, the major parasite marker of AR). Culture adapted strains await assessment of *in vitro* resistance and new parasite determinants of AR.

## Introduction

Malaria has taken a staggering historical toll on human health. However, in the 21^st^ century many areas of the world are striving for elimination and eradication. From 2010-2018 malaria cases declined globally^1^. Fast acting ACTs reduced levels of infection and death due to *Plasmodium falciparum*, the most virulent of human malaria parasites responsible for the major burden of disease. Yet substantial global levels of malaria remain. Over 400,000 deaths and 228 million world-wide cases of malaria occurred in 2018^1^. Global malaria levels failed to decrease in 2019^2^. Malaria elimination in Bangladesh (where 17.5 million people live at risk for infection) made strong progress from 2010 to 2018 reducing malaria cases by 81% (http://www.nmcp.gov.bd). Over 90% of malaria cases occur in the CHTs in three districts, Khagrachhari, Rangamati and Bandarban that border Myanmar (See Fig. 1A).

**Figure 1.**
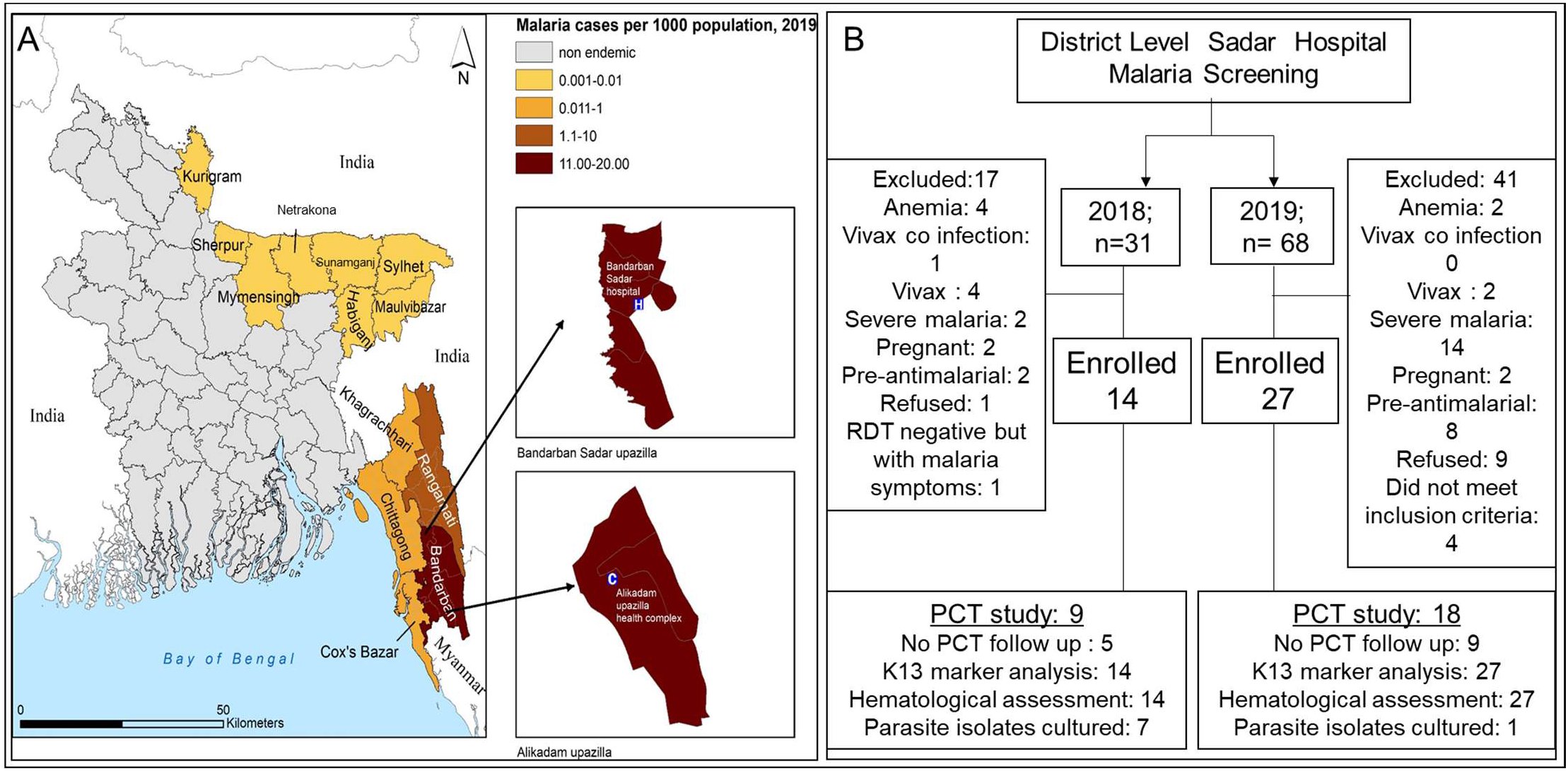
Study sites and flow chart. **A**. Map of Bangladesh highlighting the malaria endemic regions with two sub-maps showing the study sites Bandarban Sadar and Alikadam Upazilla. Data provided by the National Malaria Elimination Program Bangladesh (NMEP). **B**. Bandarban study flow chart.

Drugs are the mainstay of treating and reducing blood stages of infection that are responsible for all the symptoms and pathologies of malaria. Resistance to drugs therefore threatens malaria control. Resistance to chloroquine arose in SEA, spread through South Asia with devastating effects for malaria morbidity/mortality in Africa 30 years ago ^3^. More recent spread of resistance to front line artemisinins throughout SEA and Myanmar has led to concern that there may be spread to Bangladesh which in turn may lie in the path of global dissemination of resistance ^4 5 6^. AR may also arise independently ^7 8^, suggesting multiple ways that Bangladesh parasites may become refractory to these frontline drugs for which we still have no replacements.

Resistance to artemisinins was first reported in clinical studies as a delay in clearance of early blood stage (or ‘ring’ stage) infection from patient circulation, in response to drug treatment ^9 10^. Subsequent studies established K13 as a molecular and major, causal marker of AR in *P. falciparum* malaria^11 12^. Studies in SEA have established strong association between polymorphisms in the β-propeller domain of K13 and delayed clearance of parasites in circulation^13^. Polymorphisms in K13 and additional genes associated with resistance have been characterized by the laboratory-based ring stage survival assay (RSA^14^), an *in vitro* correlate of *in vivo* resistance. This laboratory assay is important to prove that mutations in candidate genes when engineered into sensitive parasites in local genetic or laboratory strains are causative for resistance ^15 16^ as well as strains where resistance is mediated by unknown determinants^17^. Nonetheless, the gold standard of clinical AR remains *in vivo* delayed clearance.

Detection of mutations in K13 (as well as other drug resistance markers) by application of PCR based technologies to infected blood from patients is well established and widely utilized in Bangladesh ^18^. ^19^ However, measurements of *in vivo* delayed parasite clearance in the CHTs has not been done. A major challenge is that CHTs are remote and rural (Fig. 1A). Here we present the first report of successful adaptation and establishment of the clinical assay of delayed *P. falciparum* clearance at a district level hospital in the CHTs. We also established in *vitro* cultures of Bangladesh strains isolated from the CHTs. Data collected over 2018-2019 revealed launching these capacities to fill critical gaps in monitoring AR in the CHTs and hematological parameters, in context of sharp escalation of malaria associated with climactic factors.

## Materials and Methods

### Site selection to support study design

This is an ongoing prospective study for assessing *in vivo* parasite clearance to determine presence of clinical AR and its quantitative correlation with parasitemia, comprehensive blood count parameters, AR molecular markers as well as laboratory adaptation of patient isolates to conduct further *in vitro* and genetic studies of *P. falciparum* parasite populations in Bandarban. In 2018, we established clinical capacity to measure *in vivo* AR in Bandarban Sadar Hospital. The site was chosen because it is the only hospital in the CHTs where it is possible to undertake a protocol for *in vivo* delayed *P. falciparum* clearance as recommended by the Worldwide Antimalarial Resistance Network (WWARN) ^20,21^. For this protocol WWARN recommends obtaining multiple patient blood draws over a 72 h period. The procedure therefore necessitates clinical facilities for in-patient stay for consecutive nights. This is only available at Bandarban Sadar Hospital because it is the sole district level hospital in Bandarban District that has all the required facilities.

To reach our target number of isolates for screening of K13 polymorphisms (see sample size calculation), in 2019 we added collection of parasite isolates at Alikadam Upazilla Health Complex because of its higher annual parasitological index (API; Alikadam 21.84 in 2018, 32.13 in 2019; Bandarban-0.83 in 2018; 1.6 in 2019).

### Sample size calculation

The absence of substantial information on K13 polymorphisms and no data for *in vivo* clearance in Bangladesh, limited local information on which to base power calculations. We therefore used findings from neighboring Myanmar, that reports 20% prevalence rate of the F446I mutation^22^. We assumed the same prevalence in Bandarban for all point mutations in the propeller domain of the K13 gene with 10% detected difference (precision), 80% power and 95% level of significance. On this basis, we calculated a sample size of 126 which was also expected to be within clinical capacity and existing information on patient enrollment.

### Patient screening

Patients were screened when presented with positive malaria diagnosis by microscopy and/or RDT. The inclusion criteria were *P. falciparum* mono-infection, axillary temperature ≥37.5°C or history of fever during the past 24 h, ability and willingness to comply with the study protocol for the duration of the study, informed consent from the patient or a parent or guardian in the case of children and minimum age of children ≥1 year. The exclusion criteria included severe malaria according to the WHO definition^11^ i.e. organ dysfunction, multiple convulsions, impaired consciousness. The exclusion criteria additionally included severe malnutrition, anemia (Hb <8g/dL), self-administered antimalarial or antibiotics before enrollment or pregnancy.

### Parasite clearance study procedure

Prior to administration of the first dose of Artemether-lumefantrine combination therapy (ACT) called KOMEFAN 140 (Mylan Laboratories Ltd., India), the initial parasite density and speciation was counted against 200 white blood cells (WBC; thick smear) or 2000 RBC (thin smear with duplicate slides stained with Giemsa (Merck, Germany). The smears were counted in duplicates by two independent microscopists and then averaged. Parasite density/ml was calculated by dividing the number of asexual parasites by the number of WBC counted (200) and multiplying by an assumed WBC count of 8,000/μl in thick film (or thin film, if the thick film was >250 parasites/50 WBC). Smears were considered negative when no asexual parasites were found after counting 200 WBC.

The admitted IPD (Indoor Patient Department) patients were treated by the hospital staff as per the protocol of the National Malaria Elimination Program. Briefly, one to four tablets (depending on patient’s body weight) of KOMEFAN 140 (containing 20 mg Artemether and 120 mg Lumefantrine), was administered at each of the following six time points 0 h, 8 h, 24 h, 36 h, 48 h, 60 h 72h. Blood draws were planned at 6, 12, 24, 36 and 48 h post first drug administration ^20^. Due to logistical difficulties, at the outset, the 12 and 24 h time points were respectively replaced by 18 and 36 h. However, some patients showed rapid reduction of parasites in the 12 h interval between 6 h and 18 h which limited fitting to curve. For this reason, the first four blood draws were undertaken every 6 h after administration of the first ACT (namely, 6, 12, 18, 24 h), followed by a time point every 12 h at 36, 48, 60, 72 h.

Patients were confirmed to be free of parasites and fever for at least one day and up to 72 h prior to being released from the clinic. Smears were considered negative when no asexual parasites were found after counting 200 WBC or 2000 RBCs. Patients were subsequently followed up for fever, pulse and blood pressure and malarial infection (by RDT and slides) up to 30 days at home weekly on day 9, 16, 23 and 30. No abnormalities were found. The study had no influence on the treatment decisions made by the hospital physicians.

### Analyses of parasite clearance data

Parasite clearance times (PCTs) were determined for each patient using the WWARN parasite clearance estimator (PCE) tool^21^. The parasite clearance half-life was determined by plotting serial parasite counts on the parasite clearance estimator tool by WWARN ^20,21^. The following parameters were defined: <u>Clearance rate constant:</u> the fraction by which parasite count falls per unit time. <u>Slope half-life</u>: Estimated time in hours it takes for the parasitaemia to decrease by half (50%), independent of initial parasite density. <u>PC_50, 90, 95 and 99_:</u> Estimated time in hours for parasitaemia to be reduced by 50%, 90%, 95% and 99% of its initial value (initial parasite density) respectively. All definitions above follow WWARN methodology for calculating parasite clearance ^20,21^.

### Blood cell assessment

Sysmex XP-300 Haematology Analyser (Sysmex Corporation, Kobe, Japan) was used to provide a full blood cell count including differential white blood cell counts, prior to administration of the ACT.

### Culture adaptation of patient isolates

10ml venous blood was drawn before administrating ACT from adults (≥18 years of age) and 7ml from adolescents (ages 11-17) using K_2_EDTA vacutainers (7.2 mg; 4.0 mL; Becton Dickinsonâ, Erem-bodegem, Belgium). Most of the bulk amount of parasitized venous blood committed to culture adaptation was washed, resuspended to 4% hematocrit with freshly washed uninfected red blood cells in 5ml RPMI 1640 medium containing L-glutamine, 50 mg/liter hypoxanthine, 25mM HEPES, 2g/liter glucose, 20 μg/ml gentamicin, 0.225% NaHCO_3_ and 15% human pooled plasma with 0.5% Albumax. The suspension was introduced into a culture flask placed in a candle jar (gas conditions: 5% CO_2_, 5–10% O_2_ and 85–90% N_2_) which in turn was placed in an incubator at 37° C. Culture media was changed and thin smears with Giemsa were made every day. When the parasitemia reached 2% rings, the cultures were frozen for storage. The times to reach 2% was noted. An adaptation was considered unsuccessful if no parasites emerged upto 60 days.

### DNA extraction

Genomic DNA was extracted from 200 µl whole blood using the QiaAmp blood mini kit (QIAGEN, Inc., Germany) according to the manufacturer’s instructions. 500ul of patient whole blood was cryopreserved for future experiments.

### Species confirmation by genotyping

Malaria species was confirmed by using nested PCR method following previously published method with minor modifications ^19^.

### K13 propeller domain sequence

The sequence of the propeller domain of *kelch13* gene was amplified by nested PCR adapted from previously described procedures^11 19^ An 849 bp product was amplified by ExoSAP-IT (Affymetrix). Amplified products were sequenced using the Sanger sequencing method on an ABI 3500 Genetic Analyzer (Life Technologies, USA). Sequencing primers: K13_N1_F: 5’-GCC AAG CTG CCA TTC ATT TG – 3’, K13_N1_R: 5’ – GCC TTG TTG AAA GAA GCA GA -3’. Both forward and reverse strands were aligned against *P. falciparum* 3D7 strain (PF3D7_1343700, PlasmoDB Release 46) using CLC sequence Viewer 8 (QIAGEN Aarhus A/S).

### Weather data

Weather data for 2018 and 2019 was obtained from Soil Resource Development Institute (SDRI), Bandarban, Bangladesh.

### Ethical approval

The study was approved by the Ethical Review Committee of icddr,b (Protocol # PR-17078) and the Institutional Review Board of the University of Notre Dame (Protocol # 17-10-4146).

### Maps

ArcGIS 10.4.1 (ESRI Inc., Redlands, CA) was used to generate malaria incidence map for 2019.

### Statistical analysis

Associations between two variables were assessed using Spearman correlation and Mann-Whitney analysis. All statistical analyses were performed using GraphPad Prism 7 (GraphPad Software, La Jolla, CA).

## Results

### Overall Clinical Presentation Profiles

As indicated earlier, *in vivo* parasite clearance studies in Bandarban Sadar Hospital were undertaken over two years from 2018-2019 (Fig. 1). In 2018, of 31 patients who presented with malaria symptoms, 17 were excluded because they were either infected or co-infected with *P. vivax*, showed severe malaria, were anemic or pregnant, had recent prior exposure to antimalarials, refused to consent or were rapid diagnostic test negative despite having malaria symptoms (also summarized in Fig. 1B). Of the 14 with *P. falciparum* mono-infections, 9 were enrolled in PCT studies. The remaining 5 showed initial parasitemias below 1000 parasites/µl, the threshold level for inclusion in PCT measurements. However, all 14 were subjected to K13 marker analyses and hematological assessments. In 2019, of 68 patients who presented with malaria symptoms, 27 met inclusion criteria of *P. falciparum* mono-infection without additional factors. 18 who showed initial parasitemia ≥ 1000 parasites/µl were enrolled in PCT studies (but all 27 were included in K13 and hematological analyses).

Together the data in Fig. 1 suggested that although *P. falciparum* infections were dominant, only about a third could be utilized for PCT studies due to other complications of presentations or insufficient threshold parasitemia. One unexpected observation was that the number of malaria patients presenting at the clinic increased by ∼2 fold from 2018 to 2019 (Fig. 1B). We took this in context that many levels of vector borne diseases like malaria can be affected by climate and Bangladesh is known to be vulnerable to climate change. As shown in Fig. 2A, monthly presentation of malaria patients in the clinic, closely corresponded with the monthly parasite incidence levels in the district (Spearman correlation coefficient, ρ = 0.8804, p < 0.0001) which in turn showed significant correlation to rainfall (Spearman co-efficient, ρ = 0.73, p < 0.0001 (Figure 2B), humidity (Spearman co-efficient, ρ = 0.91, p < 0.0001) and minimum temperature (Spearman co-efficient, ρ = 0.72, p = 0.0001) seen in Bandarban (Fig. 2C). This strongly suggested that the two-fold increase seen in presenting patients in 2019 was due to higher levels of malaria in the district rather than other determinants affecting access and care.

**Figure 2.**
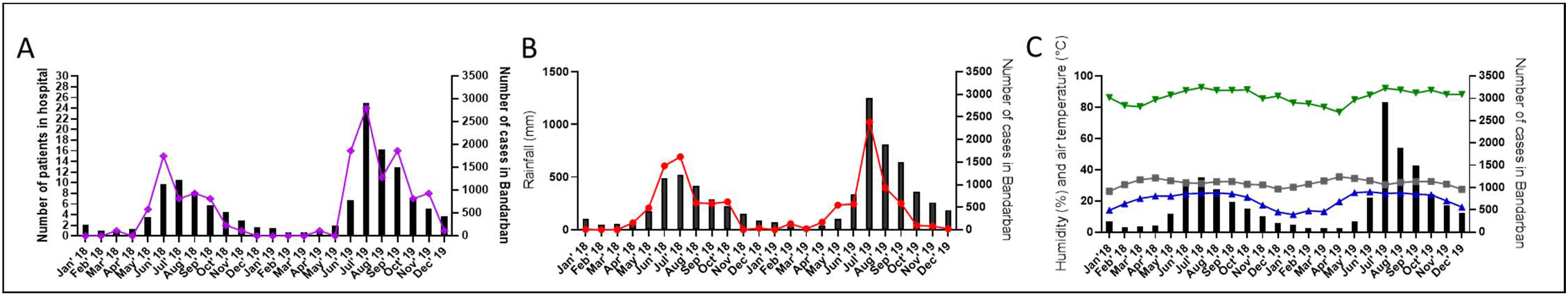
Comparative analyses of malaria patients admitted in Bandarban Sadar District Hospital, total malaria incidence and associations with climate factors in Bandarban district, 2018-2019. **A**. Total monthly malaria cases in Bandarban Sadar Hospital (purple line) and Bandarban district (bars). Spearman co-efficient, ρ = 0.8804, p < 0.0001; **B**. Total monthly malaria cases (bars) and rainfall (red line) in district, Spearman co-efficient, ρ = 0.73, p < 0.0001; **C**. Total district monthly malaria cases and maximum average temperature distribution (grey squares), Spearman co-efficient, ρ = -0.12, p = 0.58; minimum average temperature (blue triangles), Spearman co-efficient, ρ = 0.72, p = 0.0001; average humidity (green triangles), Spearman co-efficient, ρ = 0.91, p < 0.0001. Annual parasite index (API) in Bandarban Sadar was 0.83 (population at risk, n= 152,915) in 2018 and 1.60 (population at risk, n= 156738) in 2019.

Participants enrolled in PCT studies were young adults with 67% males (Table 1). At time of presentation, all but one had fever (≥ 37.5°C) and none were significantly anemic (hemoglobin < 9 g/dl for females and 10 g/dl for males) (Table 1, Supplementary Table 1 and Supporting Information SI). All participants tolerated and complied with study procedures. No serious adverse events were reported. Mild to moderate adverse events of headache, body aches, nausea, vomiting, fever or chill were observed and recorded for each participant. They were related to acute malaria illness and resolved within 72 h /3 days. Participants did not receive acetaminophen or other anti-pyretics after the first dose of the ACT KOMEFAN 140.

**Table 1:**
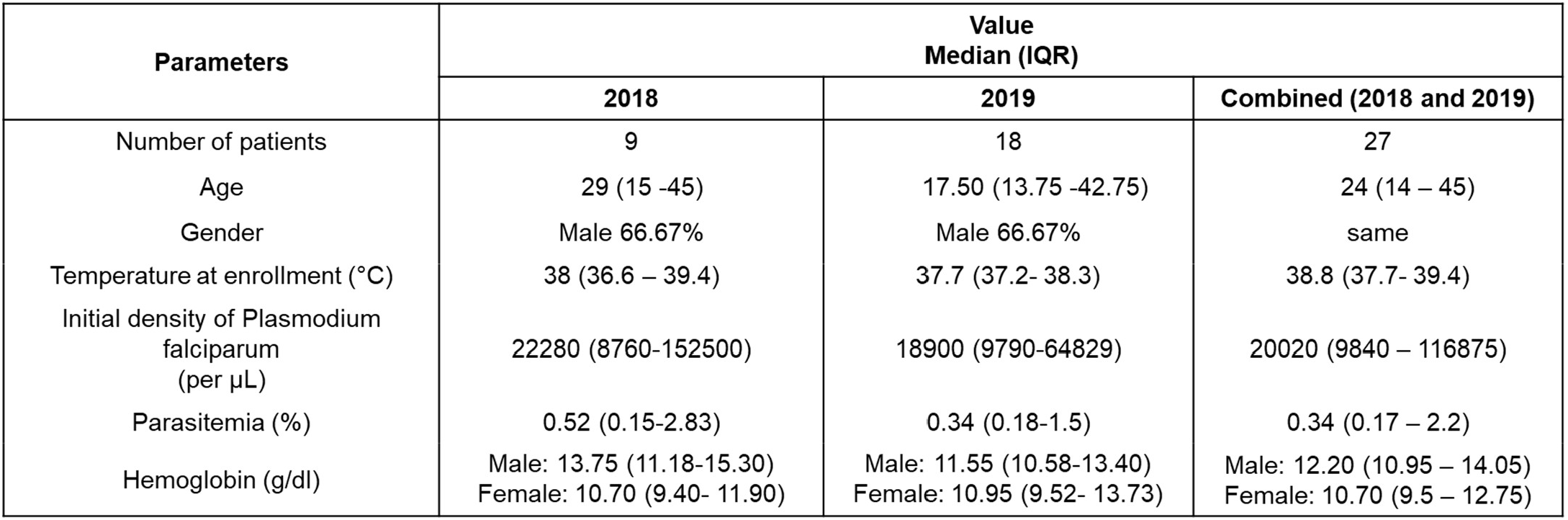
Presenting clinical characteristics and parameters of participants enrolled in PCT studies in Bandarban District Hospital Sadar, 2018 - 2019. (See also Supplementary Table 1).

| Parameters | Value<br>Median (IQR) |  |  |
| --- | --- | --- | --- |
|  | 2018 | 2019 | Combined (2018 and 2019) |
| Number of patients | 9 | 18 | 27 |
| Age | 29 (15 -45) | 17.50 (13.75 -42.75) | 24 (14 – 45) |
| Gender | Male 66.67% | Male 66.67% | same |
| Temperature at enrollment (°C) | 38 (36.6 – 39.4) | 37.7 (37.2- 38.3) | 38.8 (37.7- 39.4) |
| Initial density of Plasmodium falciparum (per µL) | 22280 (8760-152500) | 18900 (9790-64829) | 20020 (9840 – 116875) |
| Parasitemia (%) | 0.52 (0.15-2.83) | 0.34 (0.18-1.5) | 0.34 (0.17 – 2.2) |
| Hemoglobin (g/dl) | Male: 13.75 (11.18-15.30)<br>Female: 10.70 (9.40- 11.90) | Male: 11.55 (10.58-13.40)<br>Female: 10.95 (9.52- 13.73) | Male: 12.20 (10.95 – 14.05)<br>Female: 10.70 (9.5 – 12.75) |

### Clinical Outcomes and Parasite Clearance

Enrollment parasite densities were moderate in each year (Table 1). Over two years, the geometric mean was 20020/µl; 95% confidence interval 9,840 – 116,875 /µl. The median time to fever clearance was 24 with a range of (24 to 48 h; not shown). The aggregate of all data points were included in measurement of parasite clearance time independent of initial parasitemia (Fig. 3A; Table 2). Over the 27 patients, the median (range) parasite clearance half-life and time was 5.57 (1.5 – 9.6) and 24 (12–48) h, respectively. The median (range) parasite clearance half-life of patients enrolled in 2018 and 2019 was 3.1 (1.5 – 4.8) h and 6.98 (2.86 – 9.6) h respectively (Table 2; Supplemental Fig. S2A and S2B). Parasite clearance estimation profiles of individual patients with 50% clearance rates of > 5h are provided in Supplemental figure S1. Lag-phase in parasite clearance was detected in 8 out of 10 patients with delayed 50% clearance. Parasite clearance half-life based on the initial value and the time required to reach 50%, 90% and 95% (PC_50_, PC_90_, and PC_95_) are summarized in Table 2. All parasite infections were cleared by 72 h /3 days after starting treatment (Fig. 3B-C).

**Table 2:** Parasite clearance data, median (IQR) (range) of study participants in 2018 and 2019.

| Parasite clearance | 2018<br>Median (IQR) (range) | 2019<br>Median (IQR) (range) | 2018 and 2019<br>Median (IQR) (range) |
| --- | --- | --- | --- |
| Parasite clearance time (h)<sup>a</sup> | 24 (15 – 24 ) (12 – 36) | 24 (18 – 36) (12 – 48) | 24 (18 – 36) (12 – 48) |
| Parasite clearance half-life (h)<sup>b</sup> | 3.1 (1.52 – 3.09) (1.5 – 4.8) | 6.98 (3.2 – 7.89) (2.86 – 9.6) | 5.6 (2.89 – 7.38) (1.5-9.6) |
| PC<sub>50</sub> (h) | 3.56 (1.8-5.6) (0.015 – 7.98) | 3.965 (3.21-8.26) (1.25 – 13.14) | 3.56 (2.83 – 7.68) (0.015 - 13.14) |
| PC<sub>90</sub> (h) | 8 .42 (6.4-11.22) (5.32 – 13.75) | 9.45 (8.06-13.45) (5.65 – 19.22) | 9.04 (7.67 – 12.92) (5.32 - 19.22) |
| PC<sub>95</sub> (h) | 10.96 (8.23- 13.18) (6.74 – 17.14) | 12.44 (10.16-15.85) (6.87 – 21.83) | 11.83 (9.6 – 14.96) (6.74 - 21.83) |
| PC<sub>99</sub> (h) | 16.8 (12.5-18.64) (10.04 – 25.02) | 19.16 (14.63-21.4) (9.69 – 27.92) | 17.43 (14.30 – 19.74) (9.69 - 27.92) |

<sup>a</sup>First observed negative microscopy slide after first dose of ACT (hours)
<sup>b</sup>Estimated time (hours) for 50% reduction in parasite density
(Parasite clearance) PC<sub>50, 90, 95, 99</sub> (hours) = time for 50, 90, 95 and 99% reduction in initial parasite density

**Figure 3.**
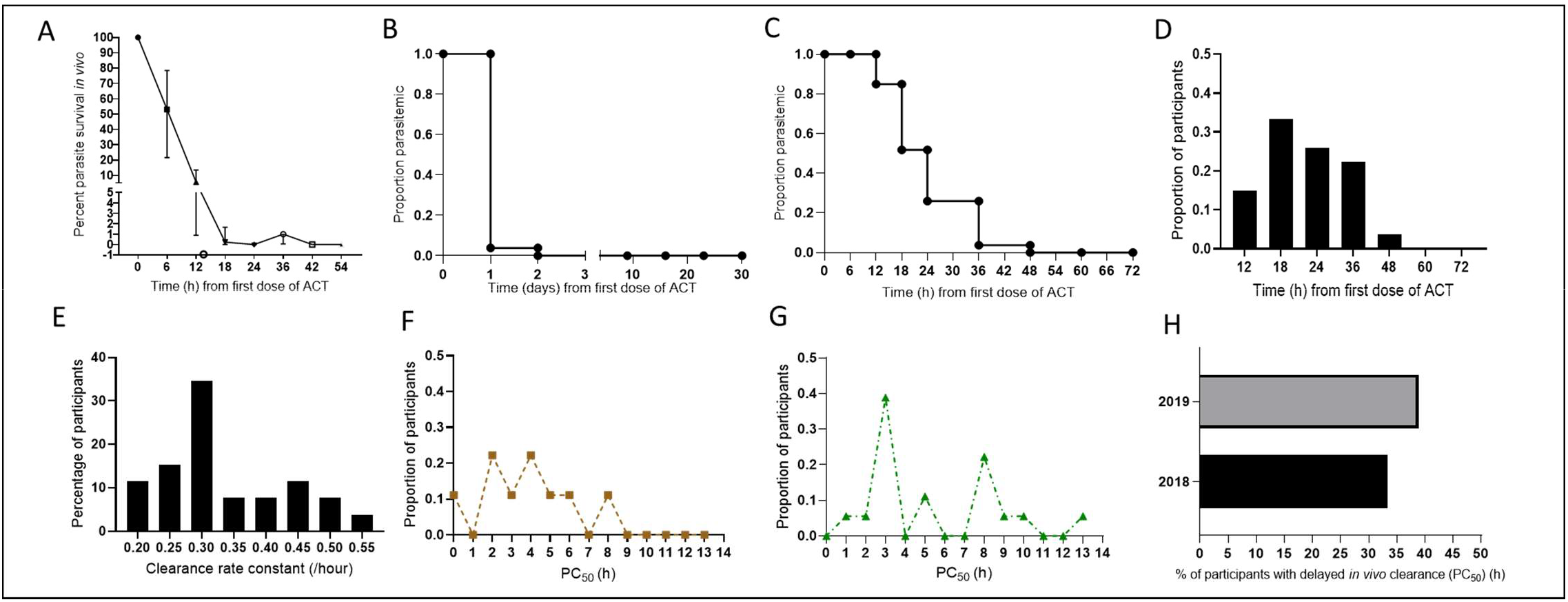
Parasite clearance data, cumulative proportion of parasitemic patients and their clearance over time, clearance rate and associated 50% clearance values. **A**. Parasite clearance curve showing linear regression of percent parasite density *in vivo* over time (hours) **B**. Cumulative proportion of parasitemic participants and their clearance over a 30 day period (follow-up on day 9, 16, 23 and 30) **C**. Cumulative proportion of parasitemic participants and their clearance over 72 hours after administration of first dose of ACT **D**. Proportion distribution of participants with time (hours) of first negative microscopy slide after administration of first dose of ACT **E**. Percentage of patients with of clearance rate constant (per hour) **F**. Proportion of participants with associated PC_50_ values, samples collected in 2018 **G**. Proportion of participants with associated PC_50_ values, samples collected in 2019 **H**. Percentage of participants with delayed *in vivo* clearance (PC_50_) in 2018 and 2019. (See also Supplementary Figure S2).

Since the number of participants were different in 2018 and 2019, we examined the proportion of participants as a function of the time in hours to a ‘slide-negative’ state to confirm that clearance was achieved by 60 h (Fig. 3C; Supplementary Fig. S2 C-D). While most patients in 2018 and 2019 reported complete parasite clearance within 48 h in 2018 and 2019; one patient reported 54h to clear all parasites, the longest time recorded for complete parasite clearance in this study (Supplemental Fig. S2 E-H). Histograms of number of participants and their clearance constant rate/hour (Fig 3D-E) suggested the highest number of participants (34%) having a clearance constant rate of 0.30/h, followed by 11% of participants having a clearance constant rate of 0.45/h, the second highest percentage of participants. Analyses of the proportion of parasites as a function of PC_50_ also supported a bimodal distribution (Fig. 3F-G) This included the expected fast clearing parasites that reach median 50% density by 3 (2.0-4.0) h, but in addition, approximately 20% of parasites showed a slower median clearance half-life of 8 (6-10) h. This was more marked in 2019 compared to 2018. The proportion of participants per year with PCT >0.5 increased from ∼ 34% to 40% from 2018 to 2019, with 38% of patients having median 50% parasite clearance of 8.4 (7.6 – 10.2) h in 2019. Together these data suggest that *in vivo* delayed clearing *P. falciparum* was reliably detected in two consecutive years, 2018 and 2019. Moreover, although the number of *P. falciparum* infection levels increased ∼2 fold, the fraction of slow clearing in 2019 was comparable to that in 2018. Nonetheless, all infections were cured, indicating that there was no failure of ACT KOMEFAN.

We subsequently undertook regression analyses to understand parasite and host factors that may influence parasite clearance. As shown in Fig. 4, there was no major association between PC_50_ and potential modifiers including age, gender and initial parasite density in either 2018 or 2019. Additionally, regression analyses failed to yield high-confidence associations between PC_50_ and initial mean corpuscular volume (MCV) mean corpuscular hemoglobin concentration (MCHC), mean corpuscular hemoglobin (MCH) white blood cells-total count (WBC/TC), mixed cell count (MXD)-monocytes– basophils–eosinophils and platelets (Fig. 5). Regression and correlation analyses, median values and range for all parameters are respectively shown in Fig. 5, and Table 3. Regression and correlation analyses between initial parasite density and hematological parameters also failed to show high-confidence associations (Supplemental Fig. S3). Together the data in Figs. 4, 5, Supplemental Fig. S3 and Table 3, suggest that initial parasitemia and blood count parameters did not make substantial contribution to *in vivo* delayed clearance phenotype. Patient hematological readouts are provided in Supplementary Table 2 and Supporting Information (SI)

**Table 3:**
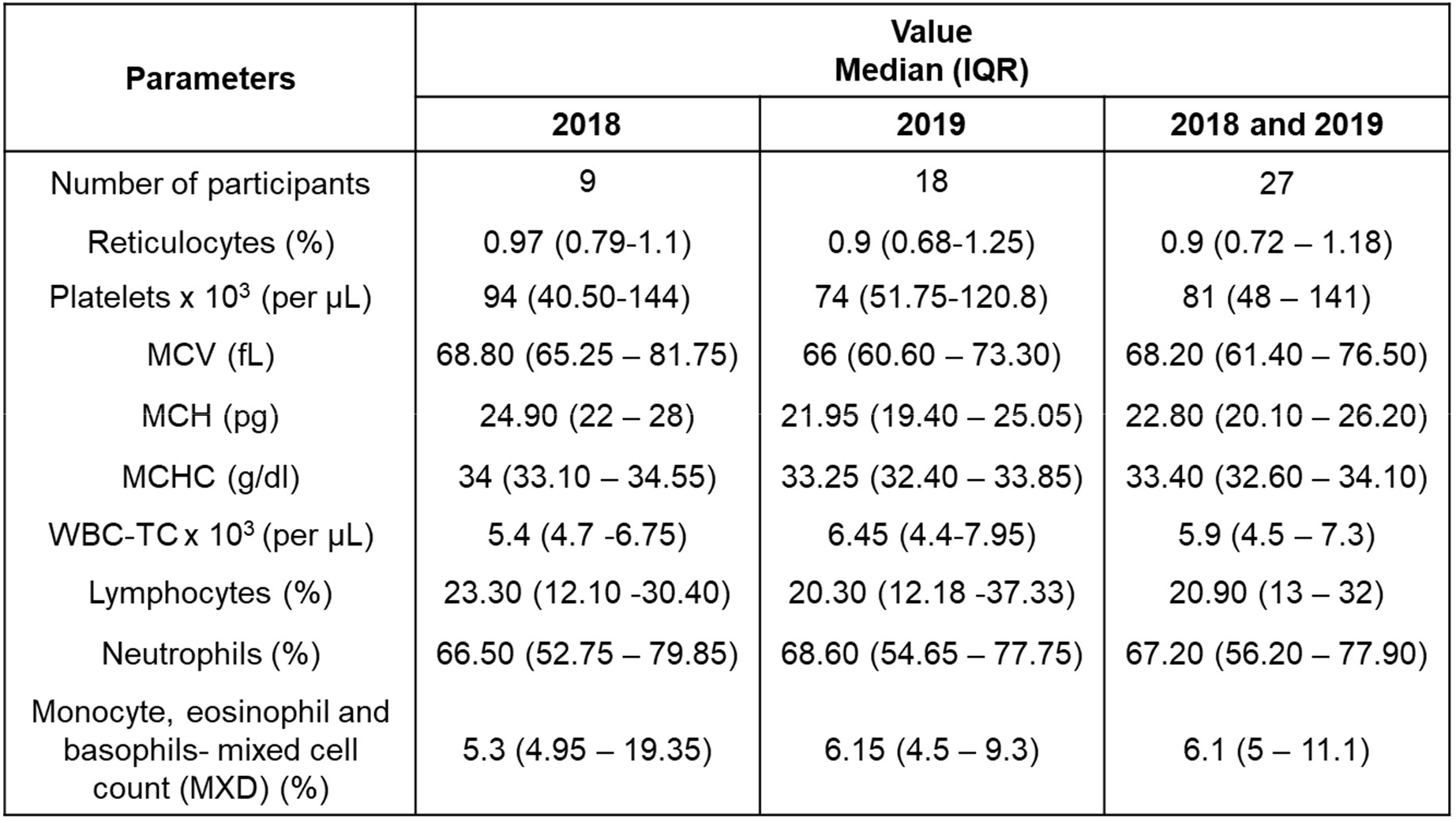
Presenting hematological and immune cell characteristics of participants enrolled in PCT studies in Bandarban District Hospital, Sadar, 2018 -2019. (See also Supplementary Table 2).

| Parameters | Value<br>Median (IQR) |  |  |
| --- | --- | --- | --- |
|  | 2018 | 2019 | 2018 and 2019 |
| Number of participants | 9 | 18 | 27 |
| Reticulocytes (%) | 0.97 (0.79-1.1) | 0.9 (0.68-1.25) | 0.9 (0.72 – 1.18) |
| Platelets x 10<sup>3</sup> (per µL) | 94 (40.50-144) | 74 (51.75-120.8) | 81 (48 – 141) |
| MCV (fL) | 68.80 (65.25 – 81.75) | 66 (60.60 – 73.30) | 68.20 (61.40 – 76.50) |
| MCH (pg) | 24.90 (22 – 28) | 21.95 (19.40 – 25.05) | 22.80 (20.10 – 26.20) |
| MCHC (g/dl) | 34 (33.10 – 34.55) | 33.25 (32.40 – 33.85) | 33.40 (32.60 – 34.10) |
| WBC-TC x 10<sup>3</sup> (per µL) | 5.4 (4.7 -6.75) | 6.45 (4.4-7.95) | 5.9 (4.5 – 7.3) |
| Lymphocytes (%) | 23.30 (12.10 -30.40) | 20.30 (12.18 -37.33) | 20.90 (13 – 32) |
| Neutrophils (%) | 66.50 (52.75 – 79.85) | 68.60 (54.65 – 77.75) | 67.20 (56.20 – 77.90) |
| Monocyte, eosinophil and basophils- mixed cell count (MXD) (%) | 5.3 (4.95 – 19.35) | 6.15 (4.5 – 9.3) | 6.1 (5 – 11.1) |

**Figure 4.**
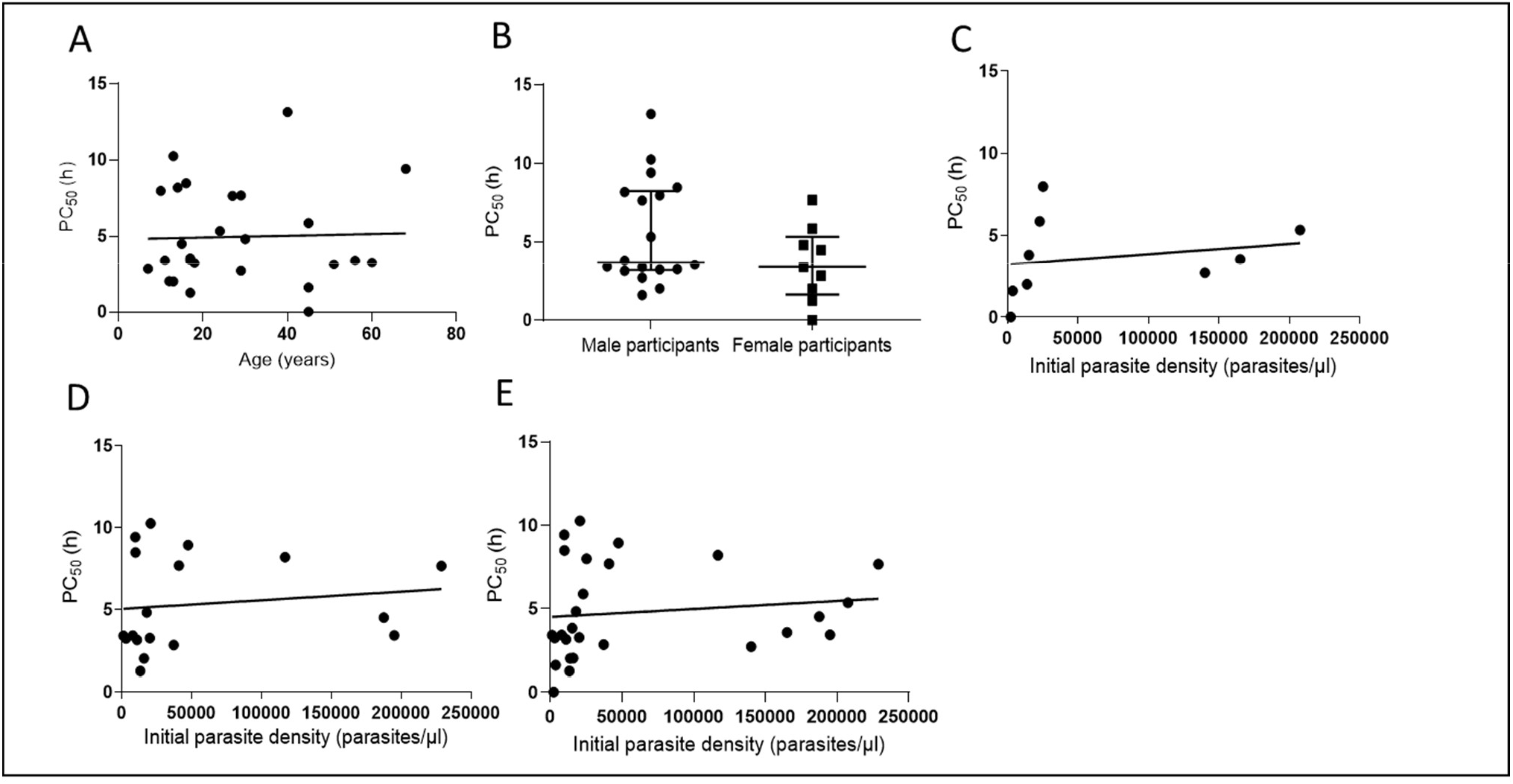
Association between 50% parasite clearance (PC_50_) and baseline parameters. **A**. Age of study participants, Spearman co-efficient, ρ = -0.026, p= 0.89, R^2^ = 0.00096 **B**. Mann-Whitney test of PC_50_ between male and female participants, p= 0.17 **C**. Initial parasite density, samples collected in 2018, Spearman co-efficient, ρ = 0.63, p= 0.076, R^2^ = 0.04 **D**. Initial parasite density, samples collected in 2018, Spearman co-efficient, ρ = 0.27, p= 0.28, R^2^ = 0.017 **E**. Initial parasite density, samples collected in 2018 and 2019, Spearman co-efficient ρ = 0.37, p= 0.054. R^2^ = 0.016

**Figure 5.**
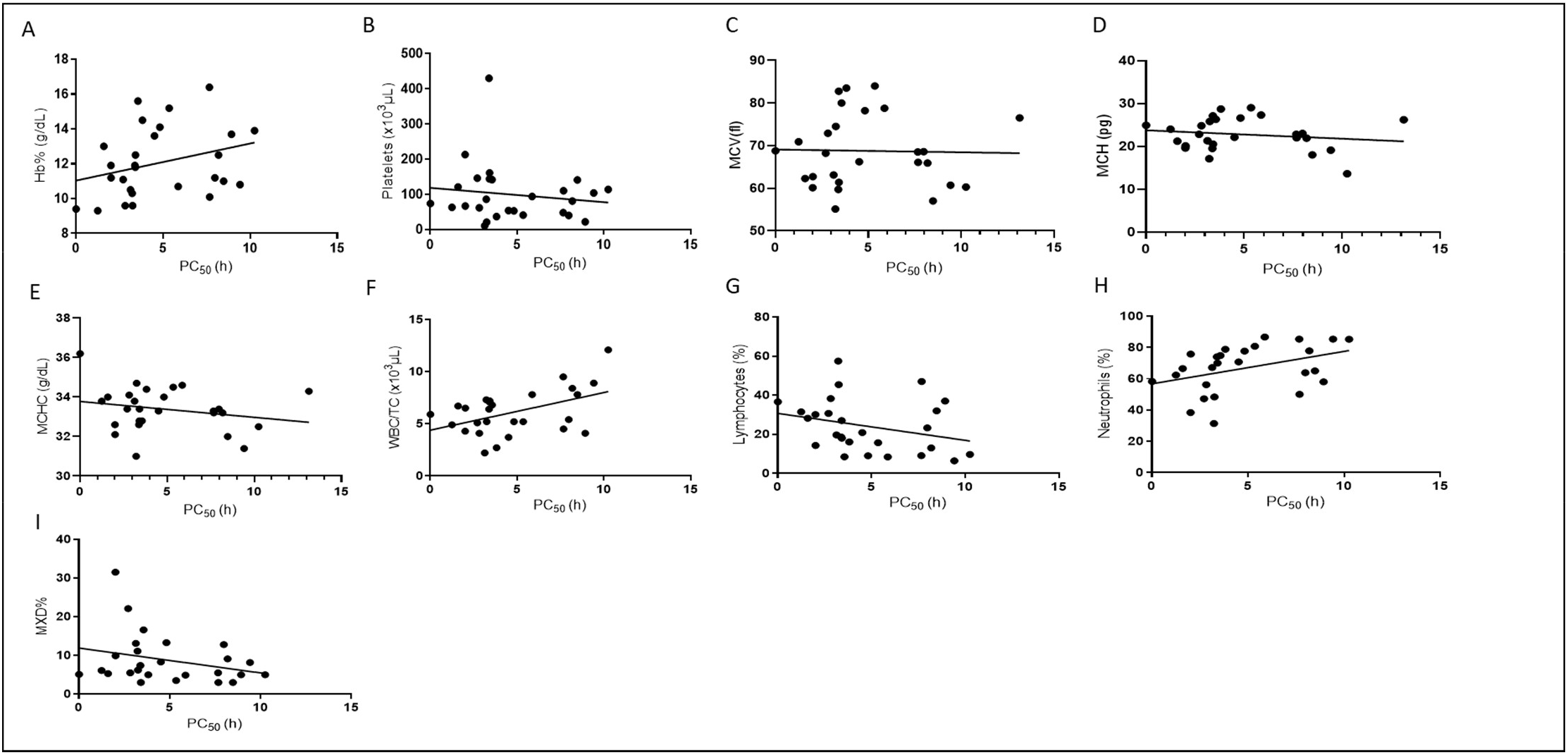
Association between 50% parasite clearance (PC_50_) and hematological parameters. **A**. Hemoglobin % (g/dL), Spearman co-efficient, ρ = 0.4, p= 0.04, R^2^ = 0.08 **B**. Platelets (µl), Spearman co-efficient, ρ = -0.13, p= 0.52, R^2^ = 0.02 **C**. Mean corpuscular volume (MCV) (fl), Spearman co-efficient, ρ = 0.03, p= 0.8, R^2^ = 0.005 **D**. Mean corpuscular hemoglobin MCH (pg), Spearman co-efficient, ρ = -0.015, p= 0.08, R^2^ = 0.03 **E**. Mean corpuscular hemoglobin concentration MCHC (g/dL), Spearman co-efficient, ρ = -0.19, p= 0.3, R^2^ = 0.05 **F**. White blood cells-total count (WBC/TC) (µl), Spearman co-efficient, ρ = 0.37, p= 0.05, R^2^ = 0.2 **G**. Lymphocytes (%), Spearman co-efficient, ρ = 0.38, p= 0.04, R^2^ = 0.08 **H**. Neutrophils (%), Spearman co-efficient, ρ = 0.48, p= 0.01, R^2^ = 0.15 **I**. Mixed cell count (MXD)-monocytes– basophils–eosinophils (%), Spearman co-efficient, ρ = -0.32, p= 0.4, R^2^ = 0.08

### K13 polymorphisms

Since comprehensive assessment of K13 polymorphisms in Bangladesh is not yet available, we used polymorphisms in neighboring Myanmar as the best guide for power calculations. F446I has been reported at 20% prevalence in Myanmar ^22^, based on which we projected requirement for collecting 126 isolates for our studies. In 2018, low numbers of patients were enrolled in Bandarban Sadar due to low levels of malaria in the area. Because of this in 2019, we added collection of parasites in Alikadam Upazilla Health Complex where the API was more than ten-fold higher than in Bandarban Sadar.

As summarized in Fig. 6 A-B, samples were collected from a total of 86 patients with acute *P. falciparum* mono-infection without severe manifestation were enrolled in Alikadam. Participants ranged from pediatric to young adults. 63% and 75% were males in 2019 and 2020 respectively. Parasite densities were moderate (2018 geometric mean, 22,460/μl; 95% confidence interval 4,860 – 35,850; 2019 geometric mean, 45,000// l; 95% confidence interval 4480-145,000) and patients were not anemic (Hb<9 g/dl). Since K13 polymorphisms associated with AR are invariably associated with β-propeller domain, we restricted analyses to this region. As shown in Fig. 6C, there were no polymorphisms identified in Bandarban Sadar. One polymorphism K13A578S was detected in Alikadam. Prior studies have reported K13A578S which is known to be a sensitive mutation, in the CHT^18^. Together these findings suggest that *in vivo* delayed clearance cannot be explained by resistance mutations in the β-propeller domain of K13.

**Figure 6.**
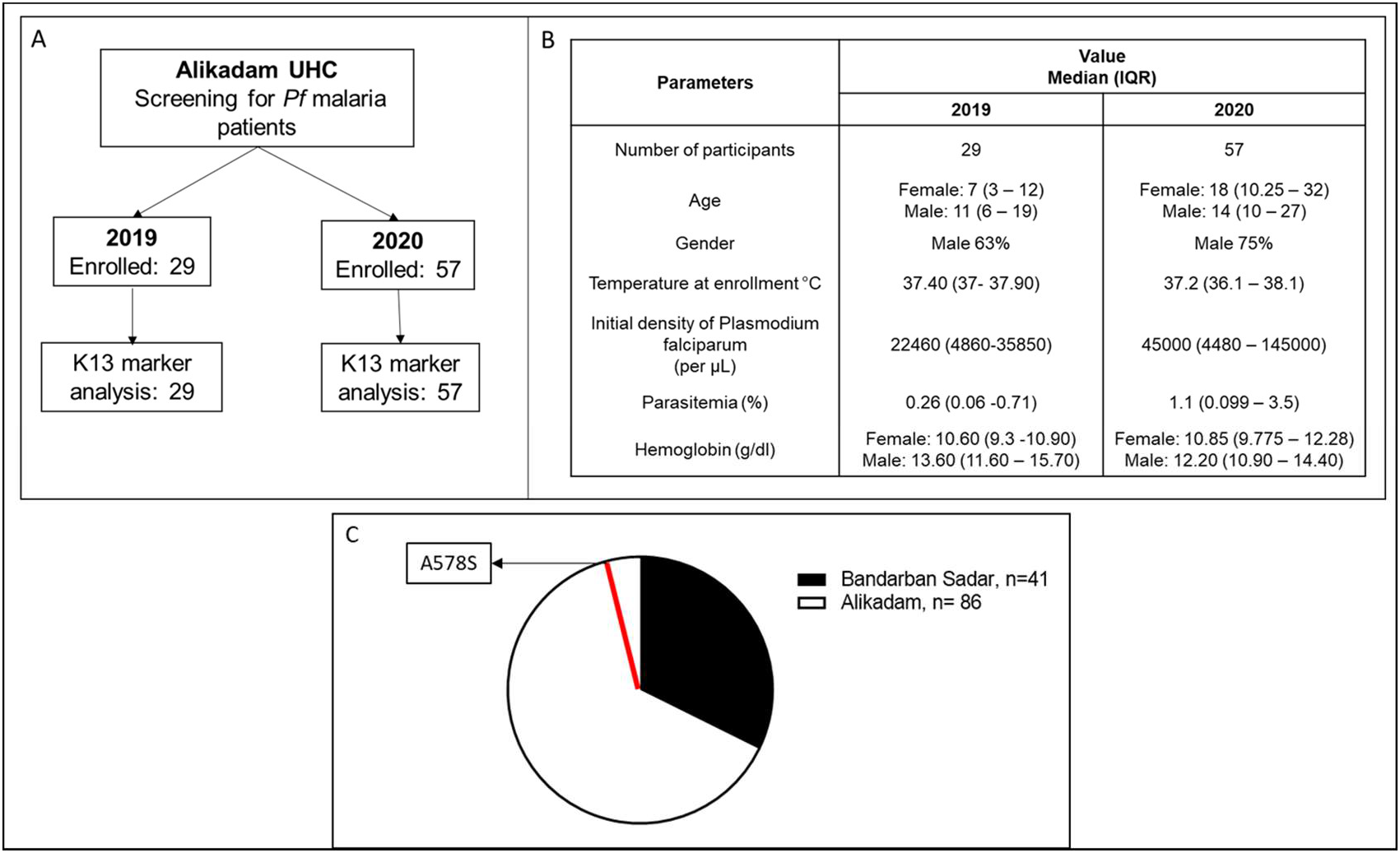
Study flow chart of Alikadam, baseline characteristics and *k13* polymorphism in all study sites. **A**. Alikadam study flow chart. **B**. Baseline characteristics, parasite clearance and incidence parameters in participants enrolled in Alikadam Upazilla Health Complex (UHC) in 2019 and 2020. **C**. *k13* polymorphisms in Bandarban Sadar and Alikadam Upazilla.

### Culture adaptation

Studies in SEA established strong association between polymorphisms in the β propeller domain of K13 and delayed clearance of parasites in circulation. However, polymorphisms in additional genes have also been associated and shown to be causal for resistance as determined in the laboratory-based ring stage survival assay (RSA) ^14 16 15^. As summarized in Fig 7A, we culture adapted 7 of 9 isolates obtained in 2018 and one of 18 in 2019. The range of initial parasitemia varied from 0.06% to 3.9%. Nonetheless 2% parasitemia was achieved within 9-12 days. In 2018, >75% of isolates were successfully adapted. However, in 2019, only one isolate (5%) could be adapted. The reasons for this variability in culture adaptation remain unclear and need to be explored in further studies.

**Figure 7.**
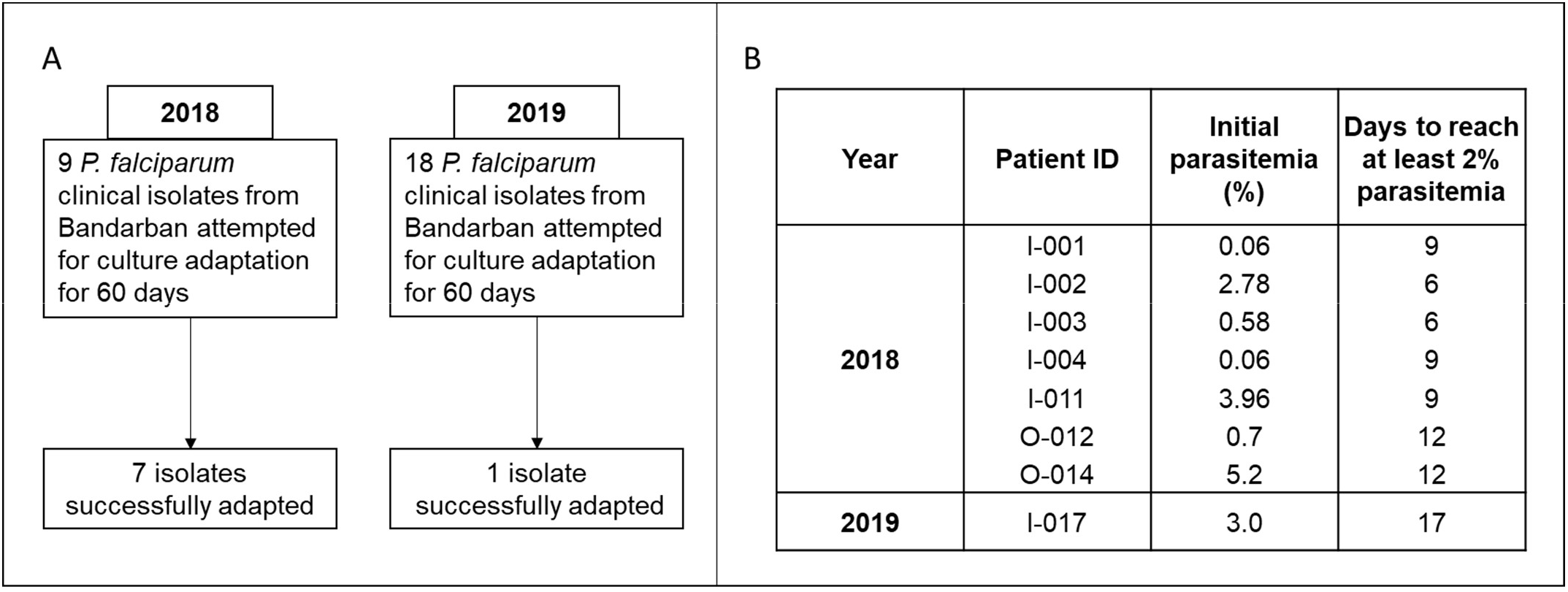
Culture adaptation. **A**. Flow chart of laboratory culture adaptation attempt in 2018 and 2019. **B**. Isolates and their initial parasitemia and days taken to reach adaptation

## Discussion

The goal of Bangladesh’s national strategic plan for malaria 2017–2021 is to eliminate the disease by 2030^23^. A key component of the plan is to prevent emergence of *P. falciparum* strains resistant to artemisinins. This needs to be accomplished in addition to interrupting local transmission, and preventing reestablishment of infection. Measuring *in vivo* delayed clearance of parasites in response to ACT treatment in malaria patients and establishment of culture-adapted Bangladesh strains to determine parasite-intrinsic resistance, are both critically important to guide strategies and interventions to achieve and sustain elimination. It will be imperative to understand whether one or both expand over time. Hence this study fills an important gap in knowledge and approaches to identifying emergence of AR in the CHTs where >90% of malaria occurs in Bangladesh.

From 2018-2019 malaria had an average of 60% resurgence across Bandarban. Infections rose in the low-API sub-district of Bandarban Sadar as well as Alikadam, with high API. The increase was well correlated with climatic factors, suggesting it may be due to increasing local transmission which is a malaria control priority that must be considered in conjunction of emerging AR^24^. Previous studies have reported on analyses of K13 polymorphisms in Bangladesh^18^. A single study^13^ reported median (range) parasite clearance half-life of 2.5 (0.7 to 5.4) in Ramu in Cox’s Bazar (where malaria incidence is low) but not in the Hill Tracts. Nor did they establish culture adaptation of Bangladesh strains, which is important, since Bangladesh strains appear to be genetically separable from SEA and Indian strains ^25^ and AR is known to be *P. falciparum* strain dependent. In our study, the clinical presentation of malaria in the Bandarban Sadar Hospital was representative of overall malaria levels and their escalation in the district, suggesting the findings have implications for the entire district and possibly the CHT. We report presentations of *P. vivax*, co-infection of *P. vivax* and *P. falciparum* as well as *P. falciparum*-induced severe disease, with substantial increase in severe disease in 2019. Consistent with prior reports ^26^ *P. falciparum* caused ∼80-90% of infections in Bandarban. However, acute, uncomplicated, mono-infection by *P. falciparum* represented 45% and 39% of patients in 2018 and 2019 respectively and only about two thirds of those had parasite densities >1000/µl.

We detected delay in ‘ring’ parasite clearance half-life > 5 h, a characteristic for AR in Bandarban. Rings are the parasite stage in the circulation and can be readily detected in a blood draw. 35-50% of enrolled patients showed delay in parasite clearance half-life for two consecutive years. Initial parasite densities > 1000 parasites /µl and time points taken every 6 hours for first 24 h after the administration of first ACT dose were critical to analyzing the data using the WWARN PCE tool. At the outset of the study, after the first draw at 6 h subsequent time points were spaced 12h apart in the first 24-30 h. But only 60% of these collections could be evaluated by the PCE tool. Upon correction to every 6 h, 100% of collections yielded interpretable data.

Across 27 patients, the median parasite clearance half-life was 5.6 h, but as many as a fifth showed much greater delayed clearance half-life of 8h. Nonetheless there was no evidence of drug failure either during the first 72h in the clinic or the following 30 days at home. This suggested that while there may be a delay in clearing rings, later stage parasites were effectively eliminated due to activity of the long lived partner drug and that the ACT is still effective. This is in contrast to Cambodia, Thailand and Vietnam where ACT combinations have lost efficacy ^6,27 28^. There was no correlation between the time to 50% of the initial parasite density (PC_50_) and initial parasitemia or blood cell counts. Notably, although platelet levels were significantly reduced relative to parasitemia (not shown) they showed no correction with PC_50_. In the case of initial Hb, lymphocytes and neutrophils, although the correlation coefficient was low (< 0.6), the p values were < 0.05, suggesting there may be weak correlation to further evaluate in subsequent studies. In prior work, correlates of immunity has been inversely correlated with delayed clearance ^29^. In future studies, measuring blood cell count every six hours during the first 24 h of parasite clearance, is expected to capture data most pertinent to the window for *in vivo* parasite delayed clearance.

A single K13 polymorphism associated with sensitive parasites was detected, suggesting that the observed *in vivo* delayed parasite clearance characteristic of AR cannot be due to a resistance mutation in K13. Further K13 resistance polymorphisms in Bandarban have not expanded to the extent (∼20%) reported in Myanmar. Establishment of parasites adapted for culture greatly facilitates understanding whether parasites that show delayed *in vivo* delayed clearance are also resistant *in vitro* (as measured by the RSA that evaluates ring stage resistance) laying the ground work to identify new parasite determinants of resistance. To the best of our knowledge this is the first report of culture adaptation of Bangladeshi parasites. A high level of success was achieved in 2018, when 7 out 9 isolates were successfully adapted. In 2019 only one (of 18) was culture adapted. The reasons remain unclear but one possibility is that because of the increase in malaria incidence, there were higher levels of long lived partner drugs in the circulation of the patient in 2019 and that compromised lab-adaptation. To address this issue, future studies need to include measurements of the concentration of partner drug after *in vivo* blood collection.

Since the Alikadam sub-district clinic facility lacks facilities for overnight stay, *in vivo* parasite clearance measurements were not undertaken there. But Alikadam was selected as a second site because it has a sub-district clinic (Alikadam Upazilla Health Complex, with 30 beds to monitor day patients) and its API was much higher than that of Bandarban (21.4 compared to 0.83 in 2018 and 32.13 compared to 1.60 in 2019). It is also closer to the border with Myanmar, where K13 resistance mutations have been reported. Work in Alikadam enabled rapid expansion of collection of infected blood samples for analyses of K13 polymorphisms and adapting Bangladeshi parasites into cultures. Building the requisite clinical capacity for overnight stay and *in vivo* parasite clearance studies will render Alikadam as a superior sub-district site for *in vivo* delayed clearance studies. This will also provide a model to expand to other sub-districts in Bandarban and the CHT, to broaden the assessment clinical AR across remote rural, forested regions with low population density, that present the major frontier of malaria in Bangladesh.

## Conclusions

We have established for the first time, capacity to measure *in vivo* parasite clearance times in response to drug treatment, a gold standard used to track AR, culture adaptation of Bangladeshi parasite strains and a model to expand both clinical and laboratory studies in remote, rural areas of Bangladesh that have the greatest concentration of malaria. We report clinical parasite clearance half-life > 5 h characteristic of AR, and culture adaptation of Bangladesh strains that empowers future laboratory analyses to determine whether clinical AR in Bandarban is an intrinsic parasite property as well as underlying new parasite determinants.

## Data Availability

All data utilized are included in the manuscript and can be found in the associated tables, figures, supplementary tables and figures

## Acknowledgments

This work was supported by the University of Notre Dame, Keough School for Global Affairs, Notre Dame Research and the Provost Office as well as National Institutes of Health USA, R01 HL 1330330 (KH). We are grateful to the study participants. We thank Dr. Mathew Sisk (Hesburgh Libraries, University of Notre Dame) for assisting in creating the incidence map of Bangladesh. We thank Dr. Arjen Dondorp (Mahidol Oxford Tropical Medicine Research Unit) for helpful discussions. We thank the Soil Resource Development Institute (SRDI), Bandarban for weather data. icddr,b acknowledges with gratitude the commitment of University of Notre Dame to its research efforts. icddr,b is also grateful to the Governments of Bangladesh, Canada, Sweden, and the UK for providing core/unrestricted support.

## Author Contributions

MKN. Conceptualization, study design, establishment and supervisor of field lab, adaptation of filed strains to *in vitro* cultures curation and analyses of all data, visualization of results, drafting and editing of manuscript.

SAS. Supervision of field lab at both the sites, in vitro culture maintenance and establishment, and data curation

AM. Conceptualization, adaptation of field strains to *in vitro* cultures, curation and analyses of all data, visualization of results, drafting and editing of manuscript.

MRHH. Supervision of field lab in Bandarban Sadar, data curation, in vitro culture maintenance, and establishment.

CSP. Clinical supervision and monitoring, and data curation

FTJ. Diagnostic PCR, K13 sequencing, and data curation.

IS. Statistical analyses and support, visualization of results and data curation.

AS. Management information systems specialist support for database support.

ASK. Study site supervision and facilitation.

RW. Conceptualization and design of blood cell count analyses, data curation and manuscript editing.

NM. Conceptualization and design of blood cell count analyses, data curation and manuscript editing.

ASM. Study supervision and facilitation (civil surgeon of Bandarban).

BC. Clinical Protocol Review and Approval University of Notre Dame.

RH. Co-investigator, study supervision

WAK. Co-investigator, study supervision

Md S. A. Field site Principal Investigator, conceptualization and project supervision, design and development of research plan, data analyses, drafting and editing of manuscript.

KH. Principal Investigator, overall conceptualization and project supervision, design and development of research plan; curation and analyses of all data, visualization of results; drafting and editing of manuscript, funding acquisition.

## Supplementary Tables and Figures

**Supplementary Table S1:**
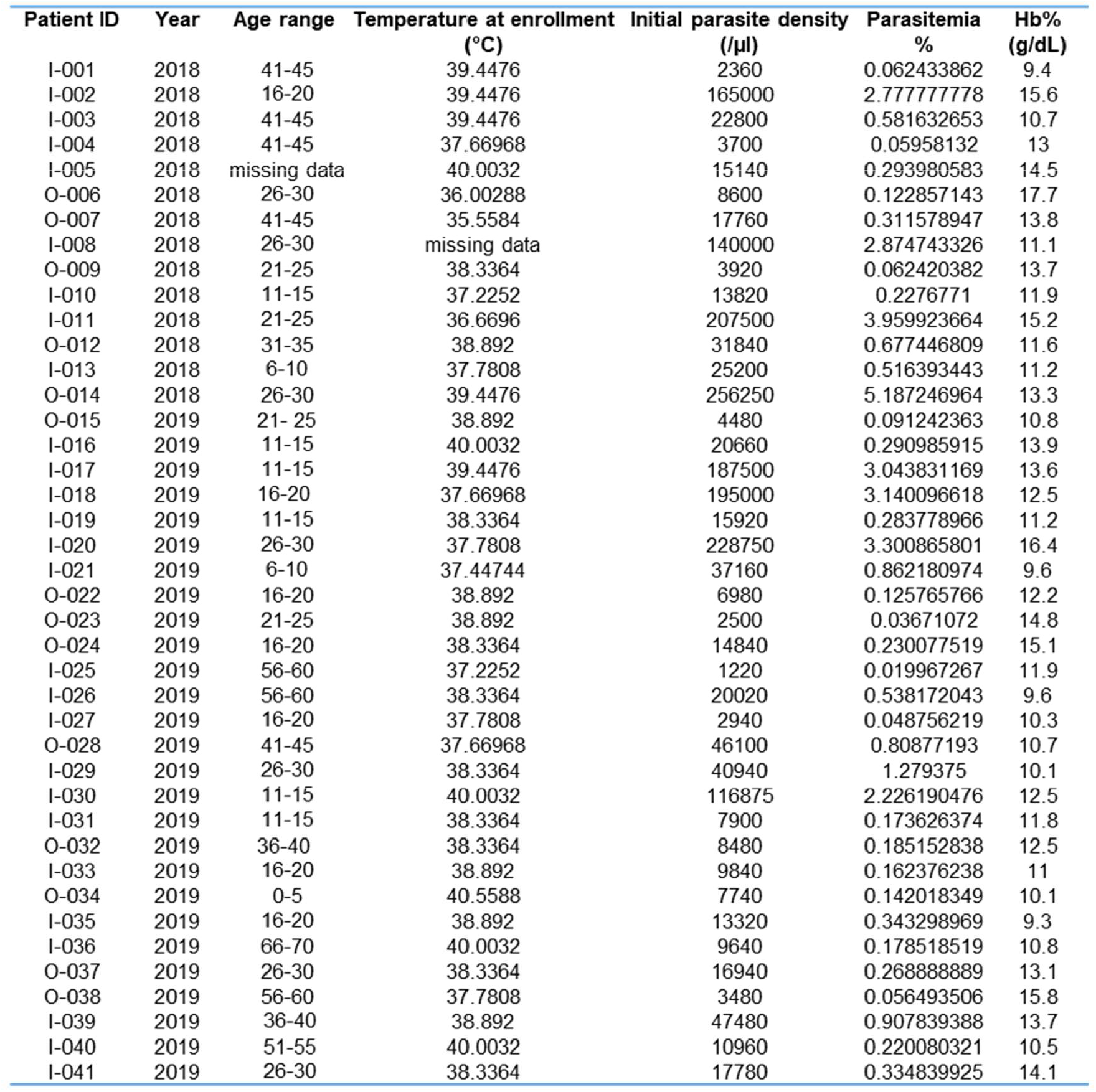
Participant age range, febrile status, parasite density (per µl), parasitemia (%) and hemoglobin (g/dl) at time of enrollment in the study in Bandarban District Hospital, Sadar, 2018 -2019.

**Supplementary Table S2:** Hematological: reticulocytes (%), platelets (per µl), mean corpuscular volume (fl), mean corpuscular hemoglobin (pg), mean corpuscular hemoglobin concentration (g/dL) and immune cell characteristics (whole blood cell total count (per µl), lymphocytes (%), neutrophils (%) and monocytes-basophils-eosinophils (%) of participants enrolled in the study in Bandarban District Hospital, Sadar, 2018 -2019.

| Patient ID | Year | Reticulocytes (%) | Platelets $\times 10^3$ ($\mu\text{L}$) | MCV (fl) | MCH (pg) | MCHC (g/dL) | WBC/TC $\times 10^3$ ($\mu\text{L}$) | Lymphocytes (%) | Neutrophils (%) | MXD (%) |
| --- | --- | --- | --- | --- | --- | --- | --- | --- | --- | --- |
| I-001 | 2018 | - | 74 | 68.8 | 24.9 | 36.2 | 5.9 | 36.6 | 58.3 | 5.1 |
| I-002 | 2018 | - | 142 | 80 | 26.3 | 32.8 | 6.8 | 8.5 | 74.9 | 16.6 |
| I-003 | 2018 | 0.9 | 94 | 78.8 | 27.3 | 34.6 | 7.8 | 8.4 | 86.7 | 4.9 |
| I-004 | 2018 | - | 121 | 62.3 | 21.2 | 34 | 6.7 | 28.2 | 66.5 | 5.3 |
| I-005 | 2018 | 1.03 | 37 | 83.5 | 28.7 | 34.4 | 2.7 | 16.1 | 78.9 | 5 |
| O-006 | 2018 | 0.84 | 84 | 75.4 | 26 | 34.5 | 7.5 | 32.2 | 48.8 | 19 |
| O-007 | 2018 | 0.79 | 165 | 75.4 | 26 | 34.5 | 5.6 | 32.2 | 48.8 | 19 |
| I-008 | 2018 | 1.03 | 146 | 68.2 | 22.8 | 33.4 | 5.1 | 30.7 | 47.2 | 22.1 |
| O-009 | 2018 | 0.9 | 86 | 72.1 | 21.8 | 30.2 | 8 | 19.8 | 67 | 13.2 |
| I-010 | 2018 | 1.3 | 213 | 60.1 | 19.6 | 32.6 | 4.3 | 30.1 | 38.4 | 31.5 |
| I-011 | 2018 | 0.88 | 41 | 84 | 29 | 34.5 | 5.2 | 15.7 | 80.8 | 3.5 |
| O-012 | 2018 | 0.68 | 46 | 73.4 | 24.7 | 33.6 | 4.8 | 47.2 | 37.5 | 15.3 |
| I-013 | 2018 | 0.52 | 40 | 68.6 | 23 | 33.4 | 5.4 | 23.3 | 63.9 | 12.8 |
| O-014 | 2018 | 0.72 | 29 | 79.8 | 26.9 | 33.8 | 2.9 | 9.7 | 83.6 | 6.7 |
| O-015 | 2019 | 0.8 | 126 | 65.8 | 21.4 | 32.5 | 5.3 | 22.9 | 65 | 12.1 |
| I-016 | 2019 | 1 | 114 | 60.3 | 13.6 | 32.5 | 12.1 | 9.7 | 85.3 | 5 |
| I-017 | 2019 | 0.6 | 54 | 66.2 | 22.1 | 33.3 | 3.7 | 20.9 | 70.8 | 8.3 |
| I-018 | 2019 | 0.4 | 161 | 61.4 | 20.5 | 33.4 | 7.2 | 18 | 0 | 0 |
| I-019 | 2019 | 1.6 | 67 | 62.7 | 20.1 | 32.1 | 6.5 | 14.3 | 75.8 | 9.9 |
| I-020 | 2019 | 0.8 | 48 | 68.5 | 22.8 | 33.3 | 9.5 | 9.1 | 85.4 | 5.5 |
| I-021 | 2019 | 0.8 | 62 | 72.9 | 24.8 | 34.1 | 4.1 | 38.3 | 56.2 | 5.5 |
| O-022 | 2019 | 0.5 | 102 | 69 | 22 | 31.9 | 4.2 | 23.3 | 55 | 21.7 |
| O-023 | 2019 | 0.6 | 181 | 67.8 | 21.7 | 32 | 9.4 | 14.7 | 79.3 | 6 |
| O-024 | 2019 | 1 | 193 | 71.1 | 23.3 | 32.8 | 5.8 | 12.5 | 79.9 | 7.6 |
| I-025 | 2019 | 0.8 | 430 | 59.7 | 19.5 | 32.6 | 6.4 | 18.6 | 74 | 7.4 |
| I-026 | 2019 | 1.8 | 21 | 74.5 | 25.8 | 34.7 | 5.2 | 45.4 | 48.4 | 6.2 |
| I-027 | 2019 | 0.6 | 86 | 55.1 | 17.1 | 31 | 7.3 | 57.5 | 31.4 | 11.1 |
| O-028 | 2019 | 0.5 | 120 | 54.8 | 17.5 | 31.9 | 7.4 | 42 | 55 | 3 |
| I-029 | 2019 | 0.7 | 110 | 66.1 | 22 | 33.2 | 4.5 | 47 | 50 | 3 |
| I-030 | 2019 | 1.40 | 81 | 65.9 | 21.9 | 33.2 | 8.4 | 13 | 77.9 | 9.1 |
| I-031 | 2019 | 0.90 | 144 | 82.8 | 27.1 | 32.8 | 7.1 | 27 | 70 | 3 |
| O-032 | 2019 | 1.10 | 121 | 79.7 | 27.3 | 34.2 | 4.8 | 34.4 | 53.7 | 11.9 |
| I-033 | 2019 | 1.40 | 141 | 57 | 18 | 32 | 7.8 | 32 | 65 | 3 |
| O-034 | 2019 | 1.20 | 476 | 59.8 | 18.5 | 31 | 14.3 | 15.7 | 77.7 | 6.6 |
| I-035 | 2019 | 1.10 | 63 | 70.9 | 24 | 33.8 | 4.9 | 31.5 | 62.4 | 6.1 |
| I-036 | 2019 | 0.60 | 104 | 60.7 | 19.1 | 31.4 | 8.9 | 6.4 | 85.4 | 8.2 |
| O-037 | 2019 | 0.80 | 51 | 61.3 | 20.8 | 33.9 | 6.4 | 38 | 62 | 0 |
| O-038 | 2019 | 1.30 | 74 | 80 | 25.6 | 32 | 5.8 | 47 | 52 | 1 |
| I-039 | 2019 | 0.90 | 22 | 76.5 | 26.2 | 34.3 | 4.1 | 37 | 58 | 5 |
| I-040 | 2019 | 1.20 | 10.5 | 63.1 | 21.3 | 33.8 | 2.2 | 19.7 | 67.2 | 13.1 |
| I-041 | 2019 | 0.90 | 53 | 78.2 | 26.6 | 34 | 5.2 | 9 | 77.7 | 13.3 |

## Supplementary Figures

**Supplementary Figure S1:**
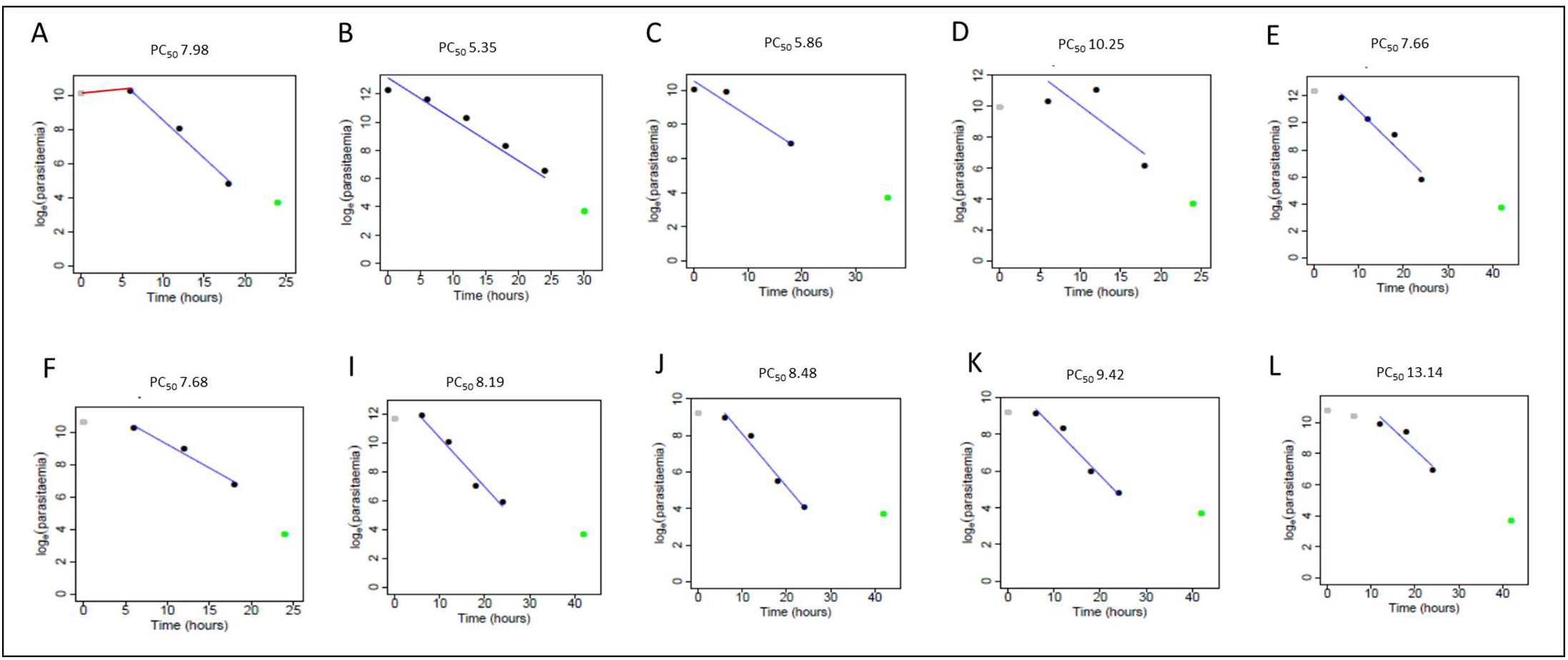
*in vivo* parasite clearance measurement in individual study participants after administration of ACT at t=0, 6, 12, 18, 24 and 36 h. At t=0, smears were made prior to drug administration. Panels **A-L** show individual WWARN plots of patients with the half-life parasite clearance exceeding 5h. Green circle: projected zero parasitemia set at log 4 to fit Tobit regression model (Flegg et al. 2011). Grey circle: Indicating a measure in lag phase, which in panel A, is shown by the red line.

**Supplementary Figure S2:**
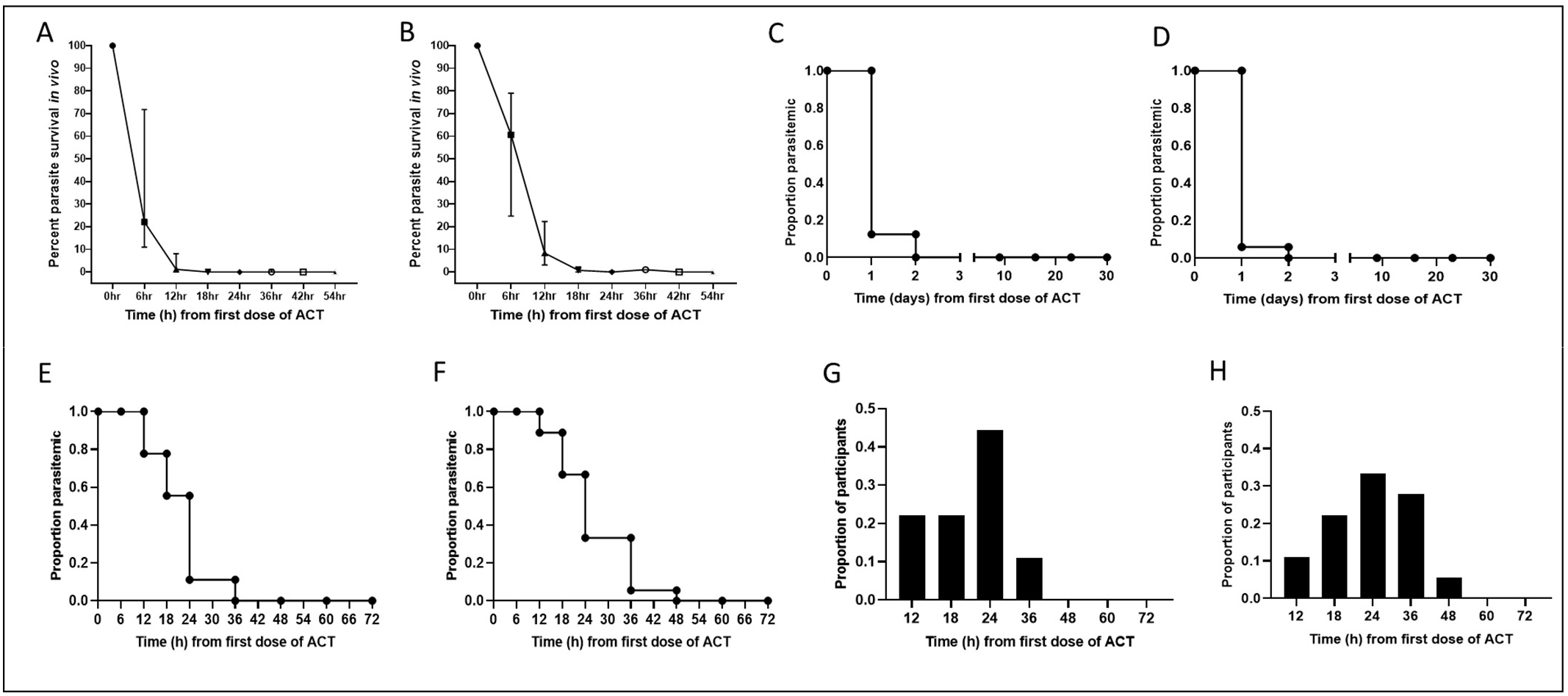
Analyses of parasite clearance data in patients enrolled in 2018 and 2019. Linear regression analyses of parasite clearance times and percent parasite density *in vivo* over time (h), **A**. 2018 and **B**. 2019. Cumulative proportion of parasitemic participants and timing of their clearance of parasites over a 30 day period (follow-up on day 9, 16, 23 and 30) **C**. 2018 and **D**. 2019. Cumulative proportion of parasitemic participants and timing of their clearance of parasites over 72 h after administration of first dose of ACT, **E**. 2018 **F**. 2019. **G-H**. Proportion of participants versus time (h) for first negative microscopy slide after administration of first dose of ACT in **G**. 2018 and **H**. 2019

**Supplementary Figure S3:**
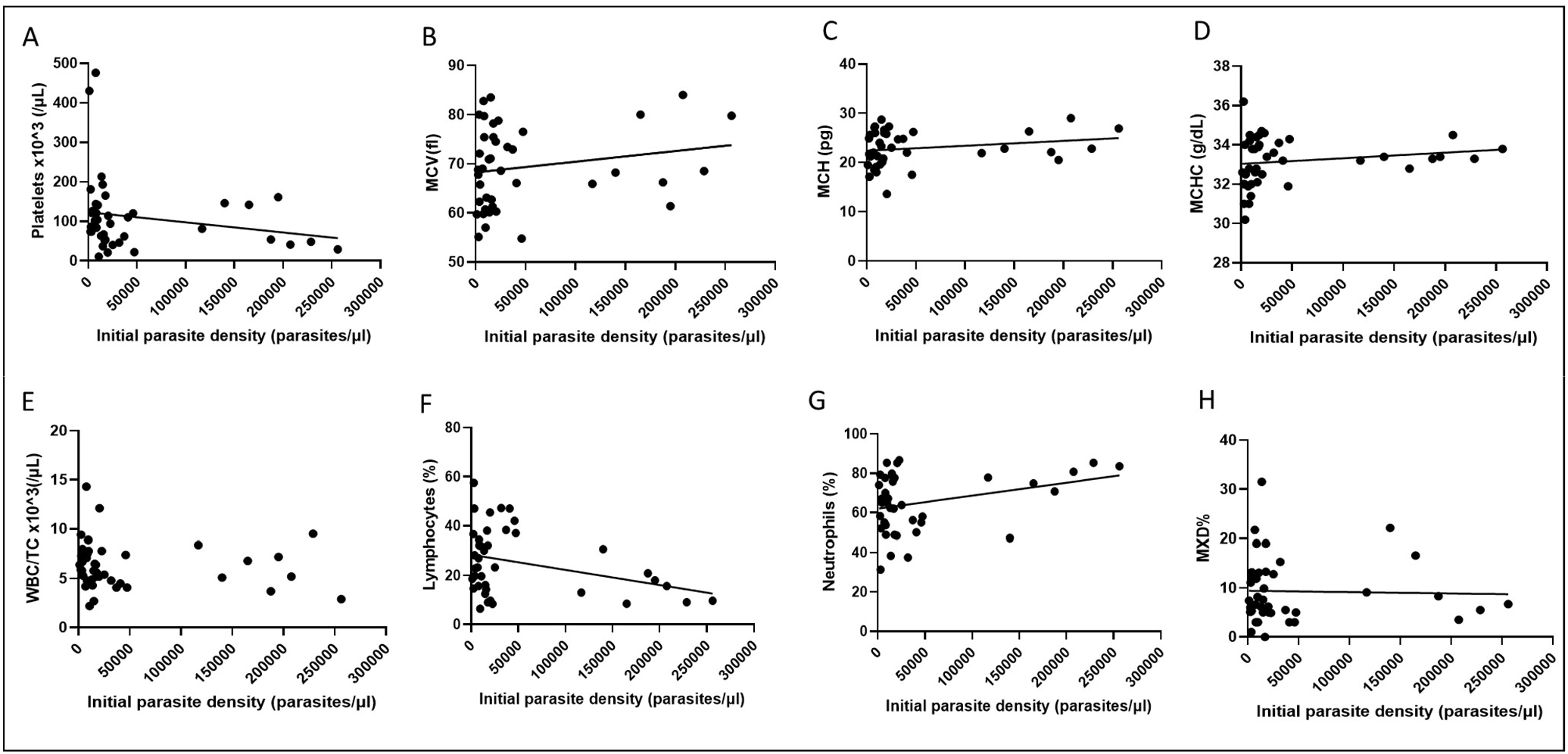
Correlation analyses between initial parasite density and hematological parameters of study participants from Bandarban District Hospital, Sadar. Linear regression and correlation analyses between initial parasite density (parasites/µl) and **A**. Platelets, Spearman co-efficient, ρ = -0.35, p= 0.02, R^2^ = 0.04. **B**. Mean corpuscular volume (MCV), Spearman co-efficient, ρ = 0.2, p= 0.2. R^2^ = 0.036 C. Mean corpuscular hemoglobin (MCH), Spearman co-efficient, ρ = 0.3, p= 0.1. R^2^ = 0.04. **D**. Mean corpuscular hemoglobin concentration (MCHC), Spearman co-efficient, ρ = 0.3, p= 0.04, R^2^ = 0.03. **E**. White blood cell-total count (WBC/TC), Spearman co-efficient, ρ = -0.2, p= 0.17, R^2^ = no fit. **F**. Lymphocytes, Spearman co-efficient, ρ = -0.2, p= 0.2, R^2^ = 0.11. **G**. Neutrophils, Spearman co-efficient, ρ = 0.17, p= 0.3, R^2^ = 0.1. **H**. Mixed cell count (MXD)-monocyte–basophil–eosinophil, Spearman co-efficient, ρ = -0.03, p= 0.8, R^2^ = 0.008

## Notes

### Competing Interest Statement

The authors have declared no competing interest.

## References Cited

1. WHO. World Malaria Report 2019. https://www.whoint/publications-detail/world-malaria-report-2019. 2019.

2. WHO. World Malaria Report 2020. https://www.whoint/publications/i/item/9789240015791. 2020.

3. Mita T, Tanabe K, Kita K. Spread and evolution of Plasmodium falciparum drug resistance. Parasitology international. 2009;58(3):201–209.

4. Imwong M, Suwannasin K, Kunasol C, et al. The spread of artemisinin-resistant Plasmodium falciparum in the Greater Mekong subregion: a molecular epidemiology observational study. The Lancet Infectious diseases. 2017;17(5):491–497.

5. Haldar K, Bhattacharjee S, Safeukui I. Drug resistance in Plasmodium. Nature reviews Microbiology. 2018;16(3):156–170.

6. Ménard D, Fidock DA. Accelerated evolution and spread of multidrug-resistant <em>Plasmodium falciparum</em> takes down the latest first-line antimalarial drug in southeast Asia. The Lancet Infectious Diseases. 2019;19(9):916–917.

7. Uwimana A, Legrand E, Stokes BH, et al. Emergence and clonal expansion of in vitro artemisinin-resistant Plasmodium falciparum kelch13 R561H mutant parasites in Rwanda. Nature medicine. 2020;26(10):1602–1608.

8. Mathieu LC, Cox H, Early AM, et al. Local emergence in Amazonia of Plasmodium falciparum k13 C580Y mutants associated with in vitro artemisinin resistance. eLife. 2020;9:e51015.

9. Noedl H, Se Y, Schaecher K, Smith BL, Socheat D, Fukuda MM. Evidence of artemisinin-resistant malaria in western Cambodia. The New England journal of medicine. 2008;359(24):2619–2620.

10. Dondorp AM, Nosten F, Yi P, Das D, Phyo AP, Tarning J. Artemisinin resistance in Plasmodium falciparum malaria. The New England journal of medicine. 2009;361.

11. Ariey F, Witkowski B, Amaratunga C, Beghain J, Langlois AC, Khim N. A molecular marker of artemisinin resistant Plasmodium falciparum malaria. Nature. 2014;505.

12. Straimer J, Gnädig NF, Witkowski B, et al. K13-propeller mutations confer artemisinin resistance in <em>Plasmodium falciparum</em> clinical isolates. Science (New York, NY). 2015;347(6220):428–431.

13. Ashley EA, Dhorda M, Fairhurst RM, Amaratunga C, Lim P, Suon S. Spread of artemisinin resistance in Plasmodium falciparum malaria. The New England journal of medicine. 2014;31.

14. Witkowski B, Amaratunga C, Khim N, et al. Novel phenotypic assays for the detection of artemisinin-resistant Plasmodium falciparum malaria in Cambodia: in-vitro and ex-vivo drug-response studies. The Lancet Infectious diseases. 2013;13.

15. Demas AR, Sharma AI, Wong W, et al. Mutations in <em>Plasmodium falciparum</em> actin-binding protein coronin confer reduced artemisinin susceptibility. Proceedings of the National Academy of Sciences of the United States of America. 2018;115(50):12799–12804.

16. Henriques G, van Schalkwyk DA, Burrow R, et al. The Mu subunit of Plasmodium falciparum clathrin-associated adaptor protein 2 modulates in vitro parasite response to artemisinin and quinine. Antimicrob Agents Chemother. 2015;59(5):2540–2547.

17. Mukherjee A, Bopp S, Magistrado P, et al. Artemisinin resistance without pfkelch13 mutations in Plasmodium falciparum isolates from Cambodia. Malaria Journal. 2017;16(1):195.

18. Mohon A, Alam MS, Bayih AG, Folefoc A, Shahinas D, Haque R. Mutations in Plasmodium falciparum K13 propeller gene from Bangladesh (2009–2013). Malar J. 2014;13.

19. Alam MS, Ley B, Nima MK, et al. Molecular analysis demonstrates high prevalence of chloroquine resistance but no evidence of artemisinin resistance in Plasmodium falciparum in the Chittagong Hill Tracts of Bangladesh. Malaria Journal. 2017;16(1):335.

20. Flegg JA, Guérin PJ, Nosten F, et al. Optimal sampling designs for estimation of Plasmodium falciparum clearance rates in patients treated with artemisinin derivatives. Malaria Journal. 2013;12(1):411.

21. Flegg JA, Guerin PJ, White NJ, Stepniewska K. Standardizing the measurement of parasite clearance in falciparum malaria: the parasite clearance estimator. Malar J. 2011;10:339.

22. Tun KM, Imwong M, Lwin KM, Win AA, Hlaing TM, Hlaing T. Spread of artemisinin-resistant Plasmodium falciparum in Myanmar: a cross-sectional survey of the K13 molecular marker. Lancet Infect Dis. 2015;15.

23. National Malaria Elimination Program. National Strategic Plan for Malaria Elimination 2017–2021. Directorate General of Health Services MoHaFW, editor Dhaka, Bangladesh: Directorate General of Health Services, Ministry of Health and Family Welfare. 2017.

24. Alam MS, Al-Amin HM, Khan WA, Haque R, Nahlen BL, Lobo NF. Preliminary Report of Pyrethroid Resistance in Anopheles vagus, an Important Malaria Vector in Bangladesh. The American journal of tropical medicine and hygiene. 2020;103(2):810–811.

25. Kumar S, Mudeppa DG, Sharma A, et al. Distinct genomic architecture of Plasmodium falciparum populations from South Asia. Molecular and Biochemical Parasitology. 2016;210(1):1–4.

26. Ahmed S, Galagan S, Scobie H, et al. Malaria hotspots drive hypoendemic transmission in the Chittagong Hill Districts of Bangladesh. PLoS One. 2013;8(8):e69713.

27. Phuc BQ, Rasmussen C, Duong TT, et al. Treatment Failure of Dihydroartemisinin/Piperaquine for Plasmodium falciparum Malaria, Vietnam. Emerging infectious diseases. 2017;23(4):715–717.

28. van der Pluijm RW, Imwong M, Chau NH, et al. Determinants of dihydroartemisinin-piperaquine treatment failure in Plasmodium falciparum malaria in Cambodia, Thailand, and Vietnam: a prospective clinical, pharmacological, and genetic study. The Lancet Infectious diseases. 2019;19(9):952–961.

29. Ataide R, Ashley EA, Powell R, et al. Host immunity to Plasmodium falciparum and the assessment of emerging artemisinin resistance in a multinational cohort. Proceedings of the National Academy of Sciences of the United States of America. 2017;114(13):3515–3520.

